# Causal Machine Learning Analysis of Radiation-Induced Leukemia and Solid Tumor Incidence in Japanese Atomic Bomb Survivors

**DOI:** 10.1101/2024.04.30.24306639

**Authors:** Igor Shuryak, Zhenqiu Liu, Eric Wang, Robert L. Ullrich, David J. Brenner

## Abstract

Uncertainty in low-dose ionizing radiation-induced health risks stems from several factors. The complex biological pathways leading to diseases like cancer are not fully understood, making it difficult to distinguish the contribution of radiation, particularly at low doses which induce only small perturbations to background disease risks. Additionally, traditional dose-response models, such as the Linear No-Threshold formalism and competing threshold or hormesis models, impose rigid assumptions on dose response shapes, causing controversy and increasing model selection uncertainty. Furthermore, these modeling strategies operate on the level of correlations/associations, and are not designed to directly address the ultimate goal of radiation epidemiology – assessing causal links between radiation and disease. A promising and rapidly-developing approach for addressing some of these challenges is causal machine learning (CML), such as double/debiased machine learning (DML), which is designed to model causal effects in multi-dimensional data sets. Our study employs DML to elucidate the causal impacts of radiation exposure on the incidence of leukemia, all solid tumors, and stomach tumors among Japanese atomic bomb survivors. Its goal was not to produce a definitive re-analysis of these data sets, but to provide a useful example of implementing CML in radiation epidemiology, which can advance the field by supplementing traditional modeling approaches. The results revealed robust positive causal effects of radiation for all three tumor types, especially for leukemia and stomach tumors. The effect magnitudes, and uncertainties, were not dramatically different at low doses than at higher doses. The influences of age at exposure, attained age, sex and other covariates on the causal effects of radiation were assessed using Shapley Additive Explanations (SHAP) values. We believe that this analysis, based on a flexible machine learning framework with a causal inference motivation and without strict dose response assumptions, provides an important contribution to radiation epidemiology.

## Introduction

While ionizing radiation is a well-known mutagen, carcinogen and cytotoxic agent, uncertainty surrounds radiation-induced health risks at low doses, stemming from several interconnected issues^1–3^. Importantly, the complex biological pathways leading to diseases like cancer or cardiovascular disease remain incompletely understood, hindering the ability to definitively attribute disease occurrence to radiation exposure^4,5^. Consequently, radiation risk estimation relies on probabilistic calculations, with population-level probabilities modeled as function of radiation dose^6^. In this situation, detecting increases in disease probability due to low radiation doses is challenging due to minimal deviations from background levels^7^.

In addition, traditional dose response modeling techniques, like the Linear No-Threshold (LNT) model, or alternative models with thresholds or hormesis, where radiation-induced risk is assumed to be zero or negative below a certain threshold dose^8^, impose rigid specific assumptions on dose-response shapes. One of the consequences of this approach is model selection uncertainty, where the researcher needs to decide which of these models to use to analyze a particular radiation effects dataset, or perhaps use a weighted ensemble of several models^9,10^. Therefore, model-specific differences in assumed dose response shapes and their implications foster controversy^11–15^.

The issue of dose response shapes, particularly at low doses, has practical significance across radiation epidemiology. For example, dose response shapes for radiation-induced cancer incidence and mortality are still not completely understood in Japanese atomic bomb survivors – a large and well-studied population which, over decades, provided a great deal of information about radiation health effects in humans^16,17^. Potential changes in dose response shape as function of dose are also relevant for other irradiated populations, such as nuclear industry workers from multiple countries investigated by the International Nuclear Workers Study (INWORKS), where “restricting the analysis to the low cumulative dose range (0-100 mGy) approximately doubled the estimate of association” between solid cancer mortality and radiation dose^4^.

These challenges make it difficult to discern radiation-induced disease effects from background “noise” and establish specific dose-response shapes, hampering current data analysis techniques^4^. Additionally, while epidemiology ultimately seeks to uncover causal relationships, such as between radiation dose and health effects, the toolkit of modeling methods that are commonly used in this field works predominantly on the level of correlation/association, rather than causation. This situation creates a need for expanding this toolkit to include newer methods which relax the strict assumptions about dose response shapes, and are specifically designed to answer causal questions.

Methods that fit these criteria are rapidly developing in the artificial intelligence (AI) / machine learning (ML) fields.^18^ Moreover, they can be readily applied to multi-dimensional data sets and handle potential nonlinear complex interactions between radiation and multiple other variables (*e.g.* demographics, genetics and lifestyle factors)^19^. Causal machine learning (CML) ^20–25^, such as double/debiased machine learning (DML) introduced by Belloni, Chernozhukov and Hansen ^26–28^, emerges as a cutting-edge field that holds significant promise for addressing the intricacies of low-dose radiation risk assessment. Contrary to the common use of ML for predictive tasks that hinge on correlations and associations—a method where distinguishing correlation from causation is notoriously difficult^29^ and conceptually distinct ^30,31^—CML focuses on directly exploring causal relationships^32^. This is of paramount importance in radiation epidemiology, where understanding whether and how radiation exposure leads to adverse health outcomes (e.g., cancer) and how these effects vary with individual characteristics (age, sex, lifestyle, genetics) is crucial^33^.

DML works by implementing three distinct models instead of one: (1) Model the causal variable/treatment (radiation dose) based on the covariates. This is a “deconfounding” operation. (2) Model the outcome (health effect) based on the covariates (but not the treatment). This is a “debiasing/denoising” operation. (3) Build a third model to relate the residuals from the first two models to each other – this relationship is interpreted as the causal effect. The causal effect can be estimated for the population (average treatment effect, ATE, or conditional average treatment effect, CATE), but also for each individual/sample. Importantly, DML has doubly robust estimator (DRE) properties – remaining unbiased even if either the treatment model or the outcome model is correctly specified, but not necessarily both – and can handle many variables in the data set.

Different types of methods can be used for models 1, 2 and 3, increasing overall power and flexibility. Essentially any type of popular ML algorithm (*e.g.* random forest^34^, XGBoost^35^, elastic net parametric regression^36^) can be used for models 1 and 2. There are also different options for model 3, including a simple linear function. For example, a powerful method for building model 3 in multidimensional data sets is causal forests (CF), which are a form of tree ensemble modeling derived from random forests^9,37,38^.

In CF, the tree splitting criterion aims to create groups of samples where the treatment effect is as constant as possible within each group. This helps identify heterogeneous treatment effects across different subgroups within the data. Additionally, CF split the dataset into two parts: one part is used to determine the tree structure (fitting the trees), while the other part is used to estimate the treatment effects based on that structure. This separation helps prevent overfitting and improves the generalization of treatment effect estimates to new data.

The main assumptions of CML methods are no unmeasured confounding (ignorability) and non-zero probability for each patient to be assigned to each treatment group (positivity). The ignorability assumption that all confounders (variables that influence both the treatment and outcome) are included in the data set usually does not hold completely, but domain knowledge about variable selection and *in silico* refutation/data manipulation methods can help to address this limitation^16^.

In this study, we employed DML to explore the causal link between radiation exposure and the occurrence of leukemia, all solid tumors, and stomach tumors among survivors of the Japanese atomic bombings. Despite their considerable potential to propel the field forward, such methodologies so far remain underutilized in radiation epidemiology^39^. We hope that this example will encourage more extensive implementation of causal inference machine learning techniques - not only DML, but potentially also generalized propensity score matching (GPS^40,41^) and targeted maximum likelihood (TML^42,43^) - to study ionizing radiation effects.

## Methods

### Data collection and pre-processing

We analyzed the following publicly available data on leukemia, all solid tumor, and stomach tumor incidence in Japanese atomic bomb survivors: (1) Incidence data set of leukemia, lymphoma and multiple myeloma among atomic bomb survivors: 1950 to 2001 follow-up. It contains 402 leukemia cases among 120,005 people with 3,842,918 migration-adjusted person years of follow-up. (2) Radiation Effects Research Foundation Life Span Study Solid Cancer Incidence Data Sets. 1958 to 1998 follow-up. It contains 18,645 cancers among 111,952 people with 2,939,361 migration-adjusted person years of follow-up. (3) Stomach tumors (4,730 cases) were analyzed separately from all solid tumors. The reason for this subset analysis was that all solid tumors represents an amalgamation of multiple tumor types with different biological mechanisms, whereas stomach tumors form an important and more uniform component of this group in the Japanese population.

The data pre-processing steps for each data set involved the removal of records with missing dose values, and recoding dose to Gy instead of mGy (creating the Dose_Gy causal variable) based on appropriate organ doses. City was coded as: 0 = Hiroshima, 1=Nagasaki; and Sex as: 0=Female, 1=Male. Variables such as "nic" indicating not in city and "ahs" representing part of the Adult Health Study clinical cohort (for solid tumors) were also encoded as binary features. Additionally, rows with more than one tumor or leukemia count were split into duplicates with one count in each row, facilitating the transformation described below. A transformed outcome variable Y=ln[(1+C)/P] was created, where C represents the tumor count (either 0 or 1) in each row, and P denotes person-years. This transformation aimed to produce an approximately normally distributed outcome variable for DML analysis. The data was then randomly split into training and testing sets (75:25) to build and evaluate the models described below.

### Double/Debiased Machine Learning (DML)

To implement DML (using Jupyter notebooks in the Python programming language version 3.10.11), we constructed three separate models on the training data: a treatment model for predicting radiation dose (Dose_Gy) as function of covariates, an outcome model for predicting transformed tumor incidence (Y) as function of covariates and excluding the treatment, and a causal forest model linking the residuals of these two models. The covariates were: age at exposure (agex), attained age (age), city, sex, nic, and ahs (for solid tumors).

Several machine learning algorithms were assessed through cross-validation (10-fold, repeated 10 times) to select which algorithm provided the best fit for the treatment and outcome models. The algorithms were: CatBoost^44^, elastic net (EN)^36^ regression, light GBM (LGBM)^45^, linear boost (LinBoost, implemented by the linear-tree Python package)^46^, linear regression (LR), random forest (RF), and XGBoost. The performance metrics (averaged over cross-validation folds and repeats) used in their comparison were: root mean squared error (RMSE), mean absolute error (MAE) and coefficient of determination (R^2^). The decision of which algorithm performed best for which model type and for which tumor type was made manually by inspecting all three performance metrics.

Based on this information, DML was constructed and applied to each tumor data set, using the EconML Python package (https://econml.azurewebsites.net/). The syntax for DML implementation was CausalForestDML(model_y=A, model_t=B, cv=10, mc_iters=10, n_estimators=200, random_state=N), which represents an instance of the CausalForestDML class from the EconML library - a machine learning model designed to estimate the causal effect of a treatment variable (Dose_Gy) on an outcome variable (Y). Here model_y is the outcome model, where A is the best-performing algorithm for the particular tumor type (*e.g.* RF). Analogously, model_t is the treatment model, where B is the best-performing algorithm for the particular tumor type (*e.g.* XGBoost). The argument cv=10 is specifying that 10-fold cross-validation should be used when training the models, mc_iters=10 is specifying that the model should perform 10 Monte Carlo iterations, n_estimators=200 is specifying that the model should use 200 trees in the causal forest, and random_state is setting the seed for the random number generator, which ensures that the results are reproducible. Sensitivity calculations were performed on the cv (5 or 10), mc_iters (5 or 10) and n_estimators (100 or 200) parameters, which found little change in the resulting causal effect estimates.

The conditional average treatment effect (CATE) estimates generated by this approach represent locally linear approximations to the dependence of the response variable Y on the treatment variable Dose_Gy. In simpler terms, the CATE is an estimate of how much the response variable Y changes for a small change in the treatment variable Dose_Gy, given the covariates X. This is analogous to the derivative of Y with respect to Dose_Gy, denoted as dY/dDose_Gy, in a local region determined by the covariates X. Therefore, the CATE estimates provide a measure of the local sensitivity of the response variable to changes in the treatment variable, taking into account the influence of the covariates. This is particularly useful in scenarios where the treatment effect may vary across different levels or groups of the covariates.

To assess their robustness and generalizability, the trained models were then applied to the separate testing dataset for each tumor type. Additionally, SHapley Additive exPlanations (SHAP)^47^ values were employed to interpret the causal forest model. SHAP values provide an importance value to each relevant feature (variable) in the model, quantifying the influence of each feature on the model’s output for any given sample. These values are particularly useful for understanding the relative impact of different features on the model’s predictions, aiding in model interpretation and insight generation. By analyzing SHAP values, one can gain deeper insights into the underlying mechanisms driving the model’s predictions and identify critical factors influencing the outcomes of interest.

## Results

### Data Set Structure

The covariates (features) used in our analysis were: agex (age at exposure), age (attained age), sex (0=female, 1=male), city (0 = Hiroshima, 1=Nagasaki), nic (not in city indicator), and ahs (adult health study indicator, present only for solid tumors and not for leukemia). Gdist (distance from the atomic bombing) was not used because it is strongly correlated with Dose_Gy (which is the causal variable), and with nic. Pearson correlations between these variables were assessed on the training data sets for all three tumor types. They were generally not strong, except for the expected strong correlation (about +0.75 on each data set) between age and agex.

### Selection of Optimal Algorithms for the Treatment and Outcome Sub-Models of DML

Results for comparing multiple ML algorithms to construct treatment-specific and outcome-specific sub-models of DML are shown in Supplementary Table 1. Random forest (RF) or XGBoost turned out to have the best performances for these models for different analyzed tumor types. The treatment/causal variable (Dose_Gy) was relatively poorly fitted based on the covariates (R^2^ < 0.5), which supports the expectation that radiation dose in this population should not be strongly predictable based on variables like age at exposure, attained age, sex, *etc*. In other words, the poor fits of the treatment models were expected and indicate that radiation dose is not “redundant” to the covariates. In comparison, the outcome variable (Y) was fitted better based on the covariates (R^2^ > 0.7), suggesting that attained age, sex *etc.* have decent (but not perfect) predictive power for cancer incidence even if radiation dose is not considered.

### DML Implementation

These sub-models were then used as the basis for fitting causal forests, which relate the sub-model residuals and interpret this relationship as the causal effect of Dose_Gy on Y. The causal effects generated here serve as locally linear approximations of how the response variable Y varies with changes in the treatment variable Dose_Gy. Essentially, the CATE quantifies the extent to which the response variable Y changes in response to slight adjustments in the treatment variable Dose_Gy, while considering the covariates X. In simpler terms, it resembles the derivative of Y concerning Dose_Gy, symbolized as dY/dDose_Gy, within a specific local region determined by the covariates X.

Therefore, these CATE estimates offer insights into the local sensitivity of the response variable to variations in the treatment variable, while considering the influence of the covariates. This aspect proves especially beneficial in scenarios where the treatment effect may differ among various levels or groups of the covariates. CATE values might also provide information about the dose-response shape. In this analysis, the tumor incidence outcome was log-transformed as described above, so a constant CATE at different dose levels indicates an upwardly-curving (convex) dose response shape.

### Causal Effects of Radiation Dose

The population-level summary of resulting causal effect estimates on training data for the three studied tumor types is shown in Table 1. For leukemia and stomach tumors, the causal effects were clearly positive and statistically different from zero, indicating that increasing radiation dose increased transformed tumor incidence in a robust manner. For all solid tumors the uncertainty was somewhat larger, with 95% CIs for the distribution of individual treatment effects slightly overlapping zero (Table 1). The likely reason is that the all solid tumors dataset is an amalgamation of multiple different solid tumor types with potentially different dose response patterns.

**Table 1.** Population summary of causal effect estimates on training data for the three studied tumor types. “Uncertainty of Mean Point Estimate” contains statistics that describe the average treatment effect (ATE). They provide a summary of the central tendency (Estimate), the uncertainty (SE, standard error), the statistical significance (z value and p-value), and the range within which the true ATE to expected lie with 95% confidence (95% CIs). “Total Variance of Point Estimate” provides similar information, but for the distribution of individual treatment effects rather than the average: their standard error and 95% CIs.

| **Leukemia:** |  |  |  |  |  |
| --- | --- | --- | --- | --- | --- |
| **Uncertainty of Mean Point Estimate** |  |  |  |  |  |
| **Estimate** | **SE** | **z** | **p-value** | **95% CIs** |  |
| 0.831 | 0.161 | 5.145 | 0.000 | 0.514 | 1.147 |
| **Total Variance of Point Estimate** |  |  |  |  |  |
|  | 0.303 |  |  | 0.287 | 1.474 |
| **Solid tumors:** |  |  |  |  |  |
| **Uncertainty of Mean Point Estimate** |  |  |  |  |  |
| **Estimate** | **SE** | **z** | **p-value** | **95% CIs** |  |
| 0.734 | 0.320 | 2.294 | 0.022 | 0.107 | 1.360 |
| **Total Variance of Point Estimate** |  |  |  |  |  |
|  | 0.659 |  |  | -0.111 | 2.368 |
| **Stomach tumors:** |  |  |  |  |  |
| **Uncertainty of Mean Point Estimate** |  |  |  |  |  |
| **Estimate** | **SE** | **z** | **p-value** | **95% CIs** |  |
| 0.906 | 0.164 | 5.524 | 0.000 | 0.584 | 1.227 |
| **Total Variance of Point Estimate** |  |  |  |  |  |
|  | 0.336 |  |  | 0.257 | 1.589 |

Individual-sample level causal effects for all analyzed tumor types as function of radiation dose are displayed in Figure 1. Each semi-transparent blue circle in Figure 1 represents a CATE estimate for an individual sample (row in the data set), with darker blue regions indicating a higher density of overlapping values. This visualization shows the almost exclusively positive nature of CATE values for the studied tumor types, particularly for leukemia and stomach tumors. This pattern persists across both training and testing datasets, demonstrating the consistency of DML estimates across different portions of the data.

**Figure 1.**
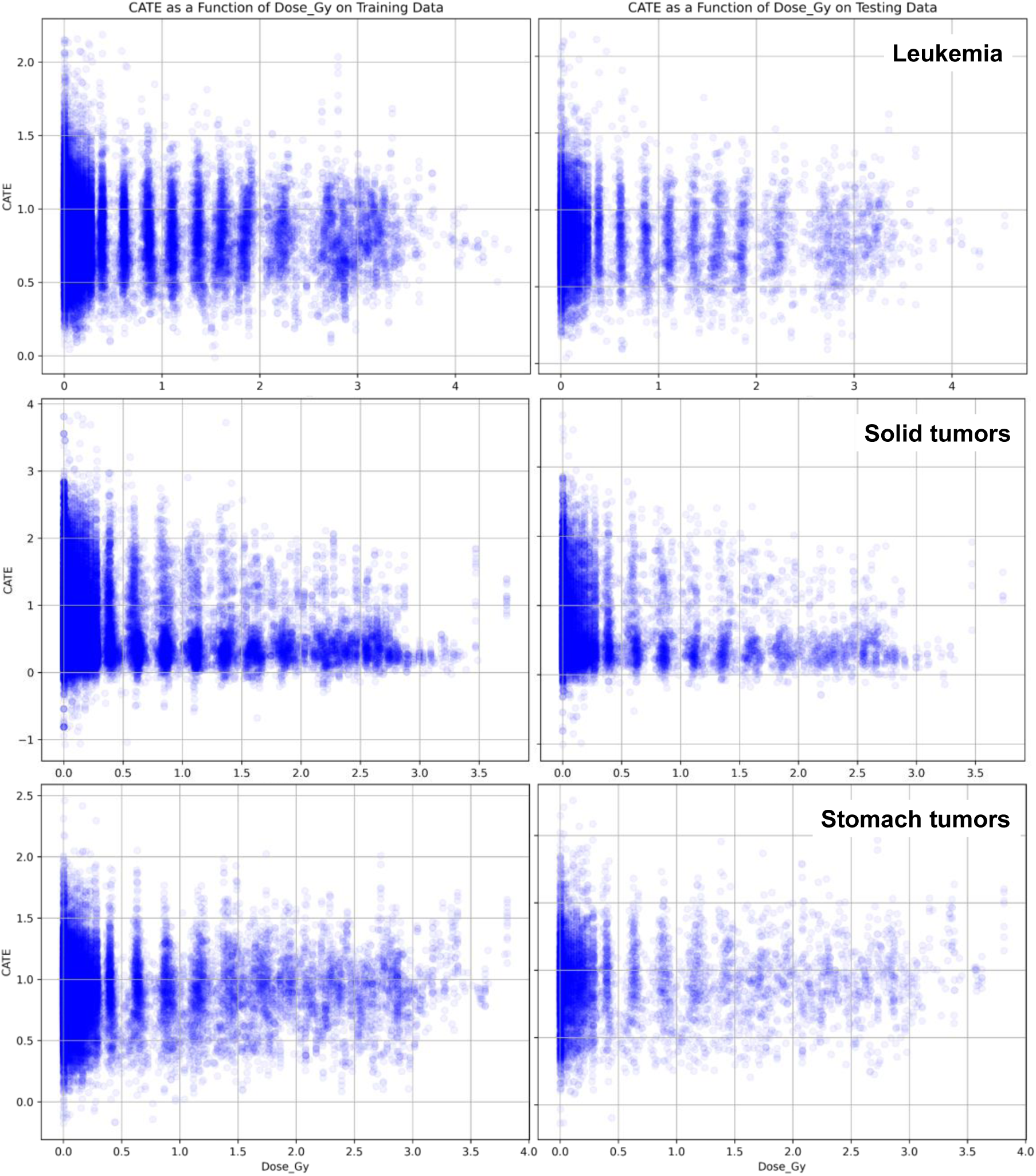
Distributions of individual-sample level CATE estimates as a function of radiation dose (Dose_Gy) on training (left) and testing (right) data for leukemia, solid tumors, and stomach tumors. Each semi-transparent blue circle is a CATE estimate for a given sample (row in the data set). Darker blue regions show where many CATE values overlap.

To better visualize how individual-sample CATE estimates varied with radiation dose, they were grouped into arbitrary dose bins (Table 2 and Figure 2). The grouping into bins was not part of the DML analysis – it was a *post hoc* procedure used for visualization only. These results showed relatively small variability in CATE estimates across different doses for all tested tumor types. For example, mean and median CATE estimates at very low doses of 0-0.01 Gy were generally similar to those at higher doses like 1-2 Gy. Notably, the uncertainties of these causal effect values (*e.g.* standard errors of the mean) were also comparable at low and high doses (Table 2 and Figure 2).

**Figure 2.**
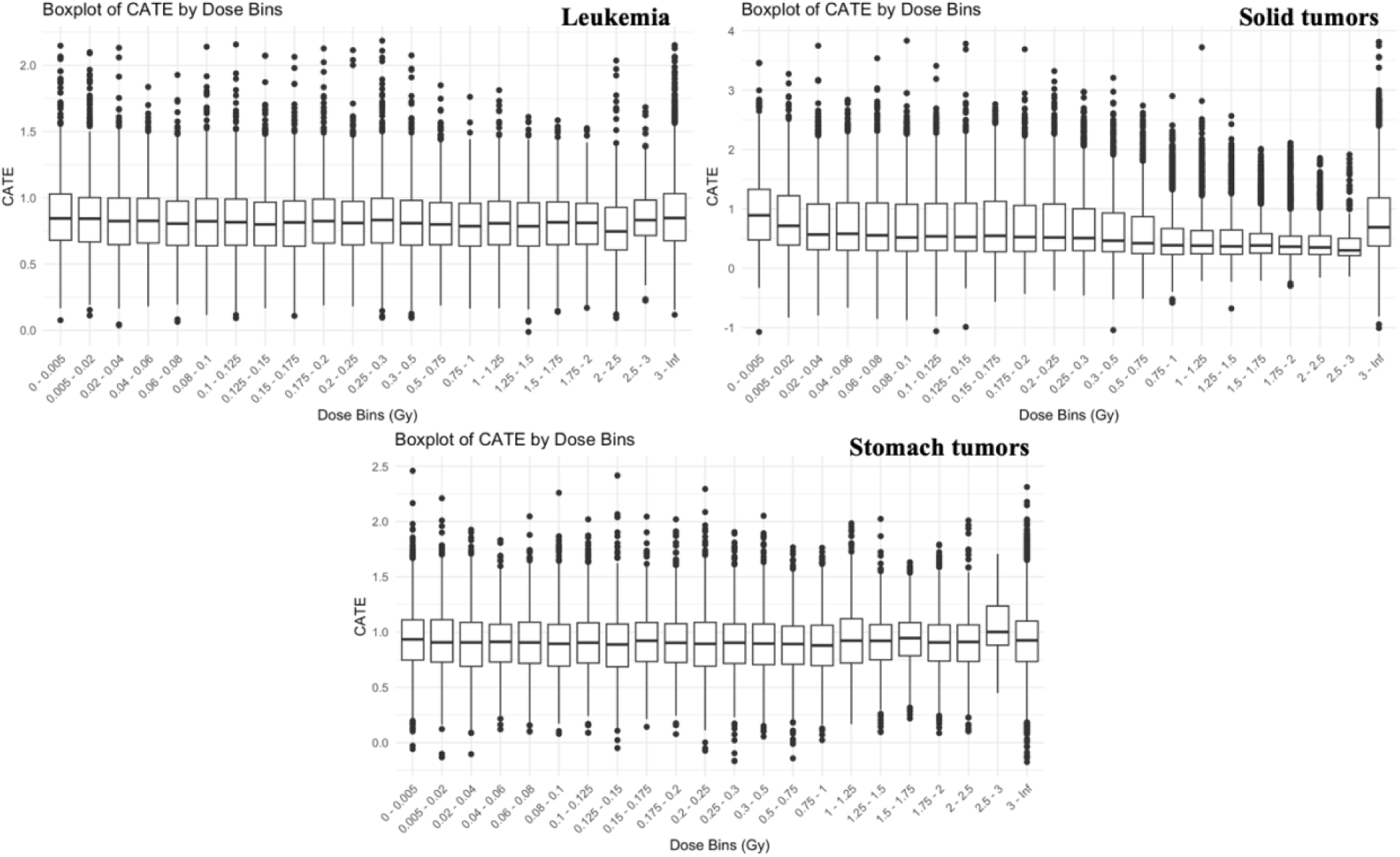
Boxplots of grouped individual-sample CATE estimates as a function of dose bins on training data for leukemia, solid tumors, and stomach tumors. As described in the Methods section, CATE was estimated by DML for each individual row/sample of the data set (shown in Figure 1), but here these CATE values were binned by dose to facilitate visualization of their potential dose dependences.

**Table 2.** Grouped individual-sample CATE estimates as a function of dose bins on training and testing data for leukemia, solid tumors, and stomach tumors. As described in the Methods section, CATE was estimated by DML for each individual row/sample of the data set (shown in Figure 1), but here these CATE values were binned by dose to facilitate visualization of their potential dose dependences. The summary statistics are: mean = average, med = median, min = minimum, max = maximum, sem = standard error of the mean. “Testing / training med” represents the ratio of median CATE values for each dose bin on testing *vs*. training data. The very high dose bins (>1 Gy) are not shown to reduce table length, but they are shown graphically in Figure 2.

| Dose bin<br>(Gy) | Leukemia |  |  |  |  |  |  |  |  |  | Testing /<br>training<br>med |
| --- | --- | --- | --- | --- | --- | --- | --- | --- | --- | --- | --- |
|  | Training data |  |  |  |  | Testing data |  |  |  |  |  |
|  | mean | med | min | max | sem | mean | med | min | max | sem |  |
| 0 - 0.005 | 0.88 | 0.87 | 0.12 | 2.15 | 0.01 | 0.83 | 0.81 | 0.17 | 1.98 | 0.01 | 0.93 |
| 0.005 - 0.02 | 0.87 | 0.86 | 0.08 | 2.15 | 0.01 | 0.84 | 0.82 | 0.20 | 2.07 | 0.01 | 0.95 |
| 0.02 - 0.04 | 0.85 | 0.84 | 0.11 | 1.97 | 0.01 | 0.85 | 0.84 | 0.21 | 2.10 | 0.01 | 1.00 |
| 0.04 - 0.06 | 0.83 | 0.83 | 0.04 | 2.13 | 0.01 | 0.82 | 0.81 | 0.04 | 1.58 | 0.01 | 0.98 |
| 0.06 - 0.08 | 0.84 | 0.82 | 0.18 | 1.84 | 0.01 | 0.83 | 0.83 | 0.18 | 1.53 | 0.01 | 1.01 |
| 0.08 - 0.1 | 0.81 | 0.81 | 0.08 | 1.73 | 0.01 | 0.81 | 0.79 | 0.06 | 1.93 | 0.01 | 0.98 |
| 0.1 - 0.125 | 0.82 | 0.82 | 0.12 | 1.92 | 0.01 | 0.82 | 0.82 | 0.17 | 2.14 | 0.01 | 1.00 |
| 0.125 - 0.15 | 0.82 | 0.81 | 0.09 | 2.16 | 0.01 | 0.84 | 0.82 | 0.26 | 1.81 | 0.01 | 1.01 |
| 0.15 - 0.175 | 0.81 | 0.80 | 0.17 | 2.07 | 0.01 | 0.82 | 0.80 | 0.18 | 1.67 | 0.01 | 1.00 |
| 0.175 - 0.2 | 0.81 | 0.81 | 0.11 | 2.06 | 0.01 | 0.82 | 0.83 | 0.20 | 1.98 | 0.01 | 1.02 |
| 0.2 - 0.25 | 0.84 | 0.83 | 0.19 | 2.13 | 0.01 | 0.82 | 0.81 | 0.30 | 1.57 | 0.01 | 0.98 |
| 0.25 - 0.3 | 0.82 | 0.80 | 0.18 | 2.11 | 0.01 | 0.84 | 0.83 | 0.33 | 1.61 | 0.01 | 1.04 |
| 0.3 - 0.5 | 0.84 | 0.83 | 0.10 | 2.19 | 0.01 | 0.84 | 0.83 | 0.15 | 2.11 | 0.01 | 1.00 |
| 0.5 - 0.75 | 0.82 | 0.80 | 0.15 | 2.07 | 0.01 | 0.82 | 0.82 | 0.09 | 1.91 | 0.01 | 1.02 |
| 0.75 - 1 | 0.82 | 0.80 | 0.24 | 1.85 | 0.01 | 0.79 | 0.78 | 0.19 | 1.66 | 0.01 | 0.98 |
|  | Solid tumors |  |  |  |  |  |  |  |  |  |  |
|  | Training data |  |  |  |  | Testing data |  |  |  |  |  |
|  | mean | med | min | max | sem | mean | med | min | max | sem |  |
| 0 - 0.005 | 0.93 | 0.94 | -0.18 | 2.18 | 0.01 | 0.90 | 0.90 | -0.03 | 2.15 | 0.01 | 0.96 |
| 0.005 - 0.02 | 0.93 | 0.94 | -0.06 | 2.46 | 0.01 | 0.91 | 0.91 | 0.12 | 1.98 | 0.02 | 0.97 |
| 0.02 - 0.04 | 0.92 | 0.91 | -0.13 | 1.96 | 0.01 | 0.91 | 0.91 | -0.13 | 2.21 | 0.02 | 1.00 |
| 0.04 - 0.06 | 0.89 | 0.90 | -0.10 | 1.93 | 0.01 | 0.91 | 0.91 | 0.21 | 1.90 | 0.02 | 1.01 |
| 0.06 - 0.08 | 0.90 | 0.91 | 0.12 | 1.83 | 0.01 | 0.90 | 0.92 | 0.16 | 1.65 | 0.02 | 1.01 |
| 0.08 - 0.1 | 0.90 | 0.90 | 0.16 | 2.05 | 0.01 | 0.90 | 0.91 | 0.10 | 1.74 | 0.02 | 1.01 |
| 0.1 - 0.125 | 0.89 | 0.88 | 0.10 | 1.86 | 0.01 | 0.90 | 0.92 | 0.08 | 2.26 | 0.02 | 1.05 |
| 0.125 - 0.15 | 0.90 | 0.91 | 0.09 | 2.02 | 0.01 | 0.90 | 0.89 | 0.09 | 1.86 | 0.02 | 0.98 |
| 0.15 - 0.175 | 0.87 | 0.88 | -0.05 | 2.42 | 0.01 | 0.91 | 0.91 | 0.14 | 2.07 | 0.02 | 1.03 |
| 0.175 - 0.2 | 0.91 | 0.92 | 0.14 | 2.04 | 0.01 | 0.94 | 0.94 | 0.21 | 1.90 | 0.02 | 1.02 |
| 0.2 - 0.25 | 0.89 | 0.90 | 0.08 | 2.02 | 0.01 | 0.93 | 0.92 | 0.25 | 1.91 | 0.02 | 1.02 |
| 0.25 - 0.3 | 0.90 | 0.89 | -0.07 | 2.29 | 0.01 | 0.89 | 0.90 | 0.11 | 2.09 | 0.02 | 1.01 |
| 0.3 - 0.5 | 0.89 | 0.89 | -0.17 | 1.90 | 0.01 | 0.93 | 0.93 | -0.10 | 1.81 | 0.02 | 1.04 |
| 0.5 - 0.75 | 0.89 | 0.90 | 0.05 | 2.05 | 0.01 | 0.90 | 0.89 | 0.24 | 1.68 | 0.02 | 0.99 |
| 0.75 - 1 | 0.87 | 0.89 | -0.14 | 1.77 | 0.01 | 0.90 | 0.90 | 0.26 | 1.73 | 0.02 | 1.01 |
|  | **Stomach tumors** |  |  |  |  |  |  |  |  |  |  |
|  | **Training data** |  |  |  |  | **Testing data** |  |  |  |  |  |
|  | **mean** | **med** | **min** | **max** | **sem** | **mean** | **med** | **min** | **max** | **sem** |  |
| 0 - 0.005 | 0.82 | 0.68 | -0.81 | 3.81 | 0.01 | 0.82 | 0.69 | -1.01 | 3.38 | 0.01 | 1.01 |
| 0.005 - 0.02 | 0.97 | 0.88 | -1.07 | 3.46 | 0.01 | 0.97 | 0.90 | -0.24 | 3.46 | 0.02 | 1.02 |
| 0.02 - 0.04 | 0.86 | 0.72 | -0.83 | 3.27 | 0.01 | 0.85 | 0.70 | -0.22 | 2.89 | 0.02 | 0.97 |
| 0.04 - 0.06 | 0.74 | 0.57 | -0.80 | 3.75 | 0.01 | 0.74 | 0.57 | -0.29 | 3.16 | 0.02 | 1.00 |
| 0.06 - 0.08 | 0.74 | 0.57 | -0.67 | 2.81 | 0.01 | 0.77 | 0.60 | -0.59 | 2.84 | 0.03 | 1.05 |
| 0.08 - 0.1 | 0.73 | 0.54 | -0.64 | 3.03 | 0.02 | 0.76 | 0.58 | -0.86 | 3.54 | 0.03 | 1.07 |
| 0.1 - 0.125 | 0.72 | 0.53 | -0.87 | 3.83 | 0.01 | 0.68 | 0.48 | -0.60 | 2.95 | 0.03 | 0.91 |
| 0.125 - 0.15 | 0.73 | 0.54 | -1.06 | 3.41 | 0.01 | 0.76 | 0.54 | -0.13 | 2.47 | 0.03 | 1.00 |
| 0.15 - 0.175 | 0.73 | 0.53 | -0.21 | 3.78 | 0.02 | 0.73 | 0.51 | -0.99 | 2.67 | 0.03 | 0.96 |
| 0.175 - 0.2 | 0.74 | 0.56 | -0.57 | 2.76 | 0.02 | 0.74 | 0.51 | -0.46 | 2.70 | 0.03 | 0.91 |
| 0.2 - 0.25 | 0.72 | 0.53 | -0.43 | 3.69 | 0.01 | 0.70 | 0.50 | -0.12 | 2.85 | 0.02 | 0.94 |
| 0.25 - 0.3 | 0.72 | 0.51 | -0.38 | 3.32 | 0.02 | 0.74 | 0.53 | -0.11 | 2.69 | 0.03 | 1.04 |
| 0.3 - 0.5 | 0.72 | 0.52 | -0.46 | 2.97 | 0.01 | 0.67 | 0.48 | -0.21 | 2.64 | 0.02 | 0.92 |
| 0.5 - 0.75 | 0.64 | 0.46 | -1.04 | 3.21 | 0.01 | 0.66 | 0.47 | -0.53 | 2.79 | 0.02 | 1.02 |
| 0.75 - 1 | 0.61 | 0.42 | -0.52 | 2.74 | 0.01 | 0.63 | 0.42 | -0.19 | 2.39 | 0.02 | 1.00 |

### Data Perturbations for Testing DML Reliability

The DML framework implemented here uses tree ensemble-based algorithms for all of its three components (RF or XGBoost for the treatment and outcome sub-models, and causal forest for the causal model component). Thus, no specific dose response shape is assumed *a priori*, and the tree ensembles should be able to reproduce even complex nonlinear dose response shapes if they exist. We conducted a series of data perturbation tests to test this in practice on the analyzed data. Specifically, we manipulated the leukemia data to generate a nonlinear dose-response curve with a derivative that changed sign within the studied dose range. This was achieved by applying the transformation Y^∗^ =Y−D−D^2^+0.3×D^4^, where Y represents the original response values, Y^∗^ are the perturbed outcome values, and D is Dose_Gy. The derivative dY^∗^/D=-1-2×D+1.2×D^3^, which changes sign from negative to positive at D=1.492. The results of DML analysis of this perturbed data set are shown in Supplementary Figure 1. They confirm that indeed the DML method generated negative CATE values at low doses, and increasingly positive values at high doses, as consistent based on the behavior of dY^∗^/D. The behavior on testing data was very similar to the one on training data.

To further validate the reliability of the DML CATE estimates as a “negative control” where no actual causal effect exists in the data, another perturbation experiment was conducted. In this case, we replaced the response variable Y with standard normal random numbers, with the expectation that any CATE estimates would center around 0. The results from this analysis (Supplementary Figure 2) corroborated our expectations. This finding was consistent across both training and testing data, further affirming the reliability of the DML method in generating CATE estimates even under simulated conditions of no actual treatment effect.

### SHAP Value Analysis

SHAP (SHapley Additive exPlanations) values are very useful for interpreting various ML model types such as tree ensemble models used here because they offer insight into the contribution of each feature to the model’s predictions. What makes SHAP values particularly valuable is their additive nature, meaning that the sum of the SHAP values for all features equals the difference between the actual model output and the average output. This property facilitates decomposing the model’s prediction for each specific sample and understanding the impact of each feature on the prediction. Additionally, SHAP values account for different combinations of features, facilitating the understanding of how interactions between features influence the model’s decision-making process.

In the context used here, SHAP values for a particular feature (covariate, like age at exposure) indicate how that feature contributed to the causal effect of radiation for the particular sample (i.e. increased or decreased the effect, and by how much). Detailed SHAP value summaries for leukemia, solid tumors, and stomach tumors are shown in Figure 3. Since CATE in this analysis represents the “slope” of the log-transformed dose response, their contribution is not on a linear scale and modest-looking changes in Figure 3 may have considerable influence on the causal effects of radiation on cancer incidence.

**Figure 3.**
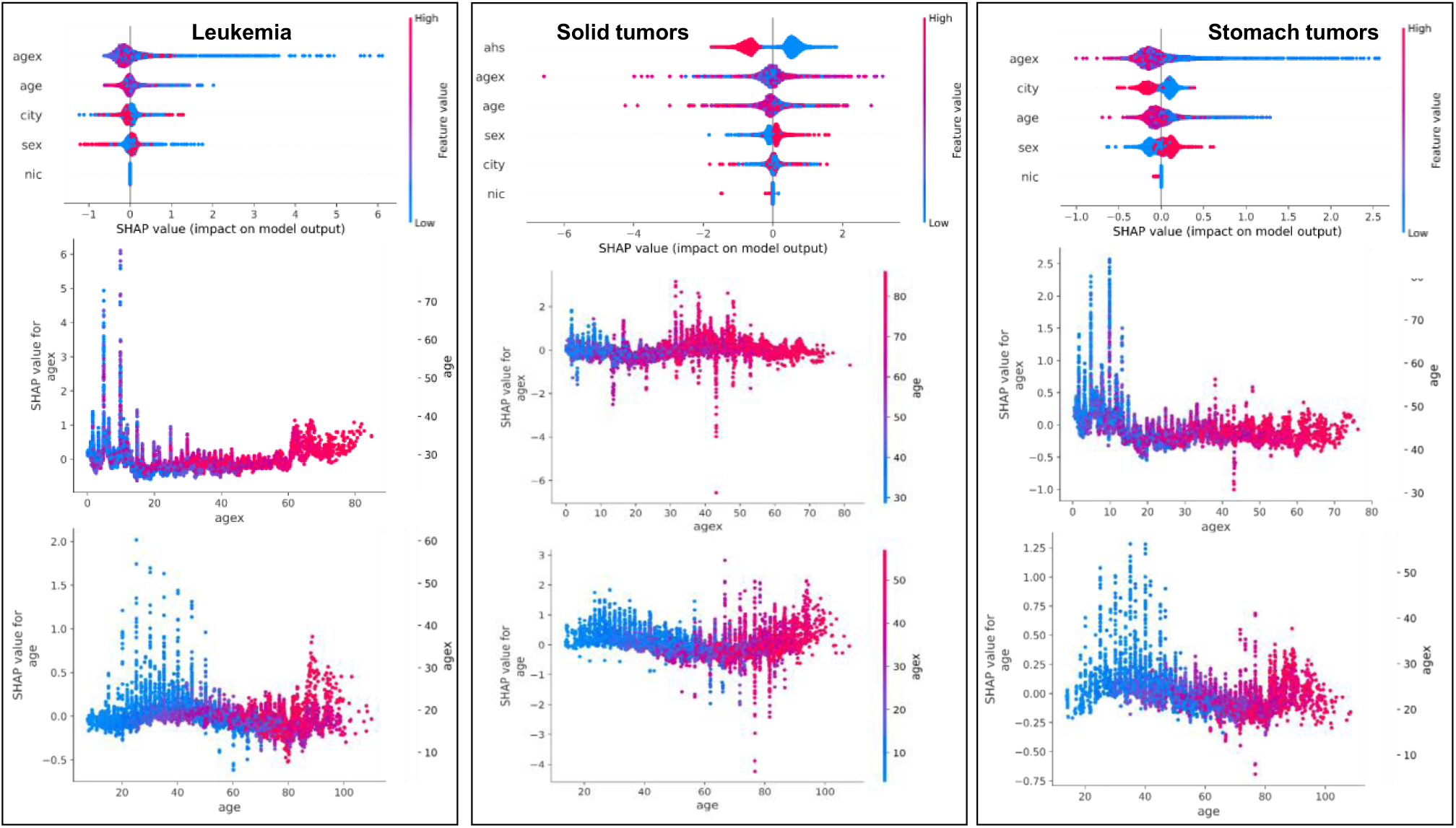
Detailed SHAP value summaries for leukemia, solid tumors, and stomach tumors. Each vertical column of three panels represents one tumor type, as labeled on top. Each circle in each panel represents the SHAP value generated by the DML model for a specific sample in the testing data set. The top horizontal row of three panels represents SHAP summary plots for the tree different studied tumor types. The features (covariates) are listed on the left side (y axis) of each panel, and the SHAP value scale is on the x axis. Positive SHAP values indicate that the feature increased the causal effect of radiation dose on the cancer outcome Y, whereas negative SHAP values indicate the opposite - that the feature decreased the causal effect of radiation. Blue circles represent low feature values, and red circles represent high feature values, according to the color scale on the right of each panel. The middle row of three panels shows a closer look at SHAP values of age at exposure (agex) as function of agex values, with the blue-red color scale indicating attained age (age). The bottom row of three panels shows a complementary closer look at SHAP values of attained age (age) as function of age values, with the blue-red color scale indicating age at exposure (agex).

For leukemia (left column of three panels in Figure 3), age at exposure (agex) and attained age (age) appear to be more important contributors than city, sex and nic (not-in city indicator). Low (young) values of agex and age tend to have positive SHAP values, suggesting a larger causal effect of radiation in younger groups, but there is variability between samples. A possible second maximum in SHAP values is visible at old agex and age. Sex does not seem to have a big effect. Correlations between SHAP values for different features were assessed using a matrix of Pearson correlation coefficients. SHAP values for age and agex were correlated +0.62, other SHAP value pairs had much lower correlations. The results suggest that most variables in the model had relatively independent contributions to the causal effect of radiation, but age and agex had correlated contributions, as could be expected since old agex cannot be associated with young age.

For all solid tumors combined (middle column of three panels in Figure 3), membership in the AHS clinical cohort (ahs) has an important influence. Age at exposure (agex) and attained age (age) appear to be more important contributors than sex, city, and nic (not-in city indicator). The direction of agex, age and sex effects has a lot of uncertainty due to variations between samples. Young agex and old age seem to have somewhat higher SHAP values, but not dramatically. SHAP values for age and agex were correlated +0.68, age and city +0.49, agex and city +0.47, agex and sex +0.45. The results suggest that age and agex had correlated contributions, as could be expected since old agex cannot be associated with young age. Other variable SHAP values were correlated more than in the leukemia analysis, suggesting that age-related, sex-related and city-related contributions in the model are inter-related. However, none of the variables came close to being completely redundant (e.g. correlations >|0.8|). Overall, these results for all solid tumors show a less clear pattern than for leukemia, likely because all solid tumors are an artificial category that includes numerous biologically distinct cancer types which are commonly grouped together in modeling analyses for convenience/statistical power, rather than for mechanistic reasons.

For stomach tumors (right column of three panels in Figure 3), age at exposure (agex), city and attained age (age) appear to be more important contributors than sex and nic (not-in city indicator). Low (young) values of agex and age tend to have positive SHAP values, suggesting a larger causal effect of radiation in younger groups – this is clearer here than for all solid tumors combined. Male sex (red values) is associated with positive SHAP values, increasing CATE. SHAP values for age and agex were expectedly correlated (+0.77), other SHAP value pairs had much lower correlations.

## Discussion and Conclusions

Biological effects of ionizing radiation are studied for over 100 years. However, several factors continue to contribute to uncertainty in estimating radiation-induced health risks, particularly at low doses. Complex biological pathways leading to diseases like cancer remain poorly understood, particularly in distinguishing radiation’s contribution at low doses, which minimally affect background disease risks. Traditional dose-response models, such as the Linear No-Threshold model, impose specific assumptions on dose-response shapes, causing controversy and increased uncertainty. These models primarily rely on correlations/associations and do not directly address the main goal of radiation epidemiology – assessing causal links between radiation and disease. Causal machine learning (CML), including double/debiased machine learning (DML), has emerged as a promising approach to address these challenges.

In this study, we used state-of-the-art CML methods to investigate the relationship between radiation exposure and incidence of leukemia, all solid tumors and stomach tumors in Japanese atomic bomb survivors. To our knowledge, these techniques remain under-utilized in radiation epidemiology, despite holding great promise for advancing the field.

The DMA-based modeling approach we implemented here did not use either parametric functions or dose bins. It used flexible tree-based ensemble models for all three components of DML, and generated conditional average treatment effect (CATE) estimates for each sample (each row in the data set). The CATE results were visualized in different ways, including being subsequently grouped into arbitrary dose bins. We also assessed how the causal effect of radiation varied as function of radiation dose, age at exposure, attained age, sex, and other covariates. The DML approach behaved reliably when translated from training to testing data, and during in data perturbation tests such as replacing the response variable with random numbers or modifying it according to a known polynomial dose response function.

The results of this study support a positive causal relationship between radiation exposure and all three studied tumor types. The estimated causal effect of radiation is very robust and positive for leukemia and stomach tumors, and somewhat less robust but still positive for all solid tumors combined. Importantly, the effect magnitudes, and also the uncertainties, were not dramatically different at low doses than at higher doses for all three tumor types. These results provide evidence against threshold or hormesis-like dose response models where the radiation effect is assumed to be zero or negative at low doses and to become positive only at higher doses. The influences of covariates such as age at exposure and attained age on these causal effects of radiation were assessed in detail using SHAP values.

The intent of this study was not to conduct a definitive analysis of Japanese atomic bomb survivor cancer incidence data, but to provide an example of using state of the art CML methods for modeling radiation effects in human populations. We believe that this example provides a useful contribution to radiation epidemiology by implementing causal inference techniques to advance the field and supplement traditional approaches that operate on the level of associations.

## Data Availability

All data produced are available online at https://www.rerf.or.jp/en/library/data-en/

## Supplementary Material

**Supplementary Table 1.** Comparative performance metrics of various machine learning algorithms for constructing the first two sub-models of DM: a treatment model for predicting radiation dose (Dose_Gy) as function of covariates, an outcome model for predicting transformed tumor incidence (Y) as function of covariates and excluding the treatment. The algorithms were: CatBoost, elastic net (EN) regression, light GBM (LGBM), linear boost (LinBoost), linear regression (LR), random forest (RF), and XGBoost. These algorithms were assessed through cross-validation (10-fold, repeated 10 times) on training data to select which algorithm provided the best fit for the treatment and outcome models for each analyzed tumor type. The performance metrics (averaged over cross-validation folds and repeats) used in this model comparison were: root mean squared error (RMSE), mean absolute error (MAE) and coefficient of determination (R^2^). For each tumor type and model type, bold font indicates which algorithm appeared to have the best performance.

| **Leukemia:** |  |  |  |  |  |  |
| --- | --- | --- | --- | --- | --- | --- |
| **Models for the outcome variable** |  |  |  | **Models for the treatment variable** |  |  |
| **Model type** | **RMSE** | **MAE** | **$R^2$** | **RMSE** | **MAE** | **$R^2$** |
| CatBoost | 1.210 | 0.939 | 0.710 | 0.801 | 0.609 | 0.199 |
| EN | 2.126 | 1.700 | 0.106 | 0.877 | 0.682 | 0.040 |
| LGBM | 1.247 | 0.976 | 0.693 | 0.805 | 0.616 | 0.190 |
| LinBoost | 1.413 | 1.115 | 0.605 | 0.840 | 0.636 | 0.119 |
| LR | 2.123 | 1.694 | 0.109 | 0.876 | 0.681 | 0.041 |
| RF | 1.130 | 0.848 | 0.748 | **0.698** | **0.469** | **0.391** |
| XGBoost | **1.113** | **0.863** | **0.755** | 0.729 | 0.546 | 0.337 |
| **Solid tumors:** |  |  |  |  |  |  |
| **Models for the outcome variable** |  |  |  | **Models for the treatment variable** |  |  |
| **Model type** | **RMSE** | **MAE** | **$R^2$** | **RMSE** | **MAE** | **$R^2$** |

| CatBoost | 0.923 | 0.696 | 0.767 | 0.584 | 0.394 | 0.255 |
| --- | --- | --- | --- | --- | --- | --- |
| EN | 1.768 | 1.354 | 0.147 | 0.632 | 0.449 | 0.126 |
| LGBM | 0.978 | 0.739 | 0.739 | 0.593 | 0.408 | 0.231 |
| LinBoost | 1.207 | 0.927 | 0.602 | 0.621 | 0.434 | 0.157 |
| LR | 1.767 | 1.354 | 0.147 | 0.631 | 0.445 | 0.129 |
| RF | **0.827** | **0.537** | **0.813** | **0.517** | **0.307** | **0.416** |
| XGBoost | 0.867 | 0.646 | 0.798 | 0.525 | 0.351 | 0.396 |
| **Stomach tumors:** |  |  |  |  |  |  |
| **Models for the outcome variable** |  |  |  | **Models for the treatment variable** |  |  |
| **Model type** | **RMSE** | **MAE** | **R<sup>2</sup>** | **RMSE** | **MAE** | **R<sup>2</sup>** |
| CatBoost | 1.107 | 0.856 | 0.747 | 0.724 | 0.538 | 0.262 |
| EN | 2.084 | 1.668 | 0.103 | 0.821 | 0.628 | 0.051 |
| LGBM | 1.162 | 0.907 | 0.721 | 0.732 | 0.546 | 0.246 |
| LinBoost | 1.313 | 1.030 | 0.644 | 0.772 | 0.573 | 0.161 |
| LR | 2.082 | 1.664 | 0.105 | 0.817 | 0.623 | 0.058 |
| RF | 1.026 | 0.737 | 0.783 | **0.615** | **0.409** | **0.466** |
| XGBoost | **1.017** | **0.787** | **0.786** | 0.643 | 0.467 | 0.418 |

**Supplementary Figure 1.**
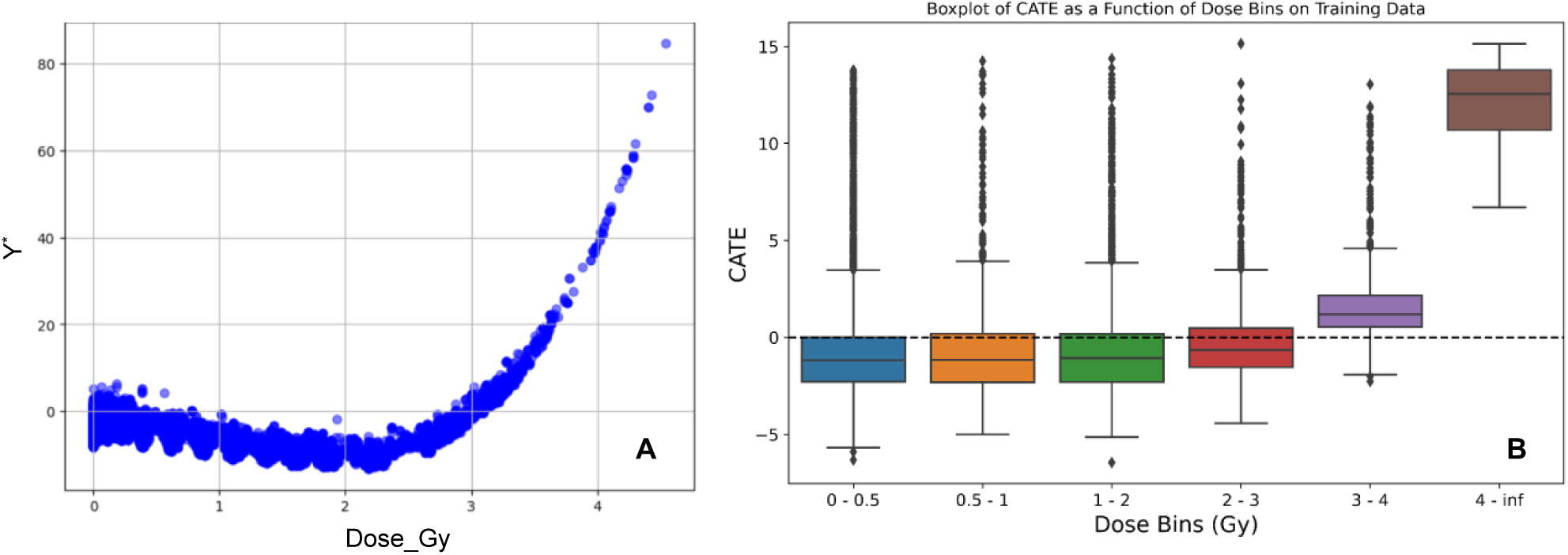
Results of DML analysis of artificially perturbed leukemia data to test the ability of the DML framework to capture complex dose response shapes. This was achieved by applying the transformation Y^∗^ =Y−D−D^2^+0.3×D^4^, where Y represents the original leukemia response values, Y^∗^ are the perturbed outcome values, and D is Dose_Gy. Y^∗^ values are plotted in panel A. A boxplot of individual-sample CATE estimates generated on this perturbed data set, grouped into arbitrary dose bins, is shown in panel B.

**Supplementary Figure 2.**
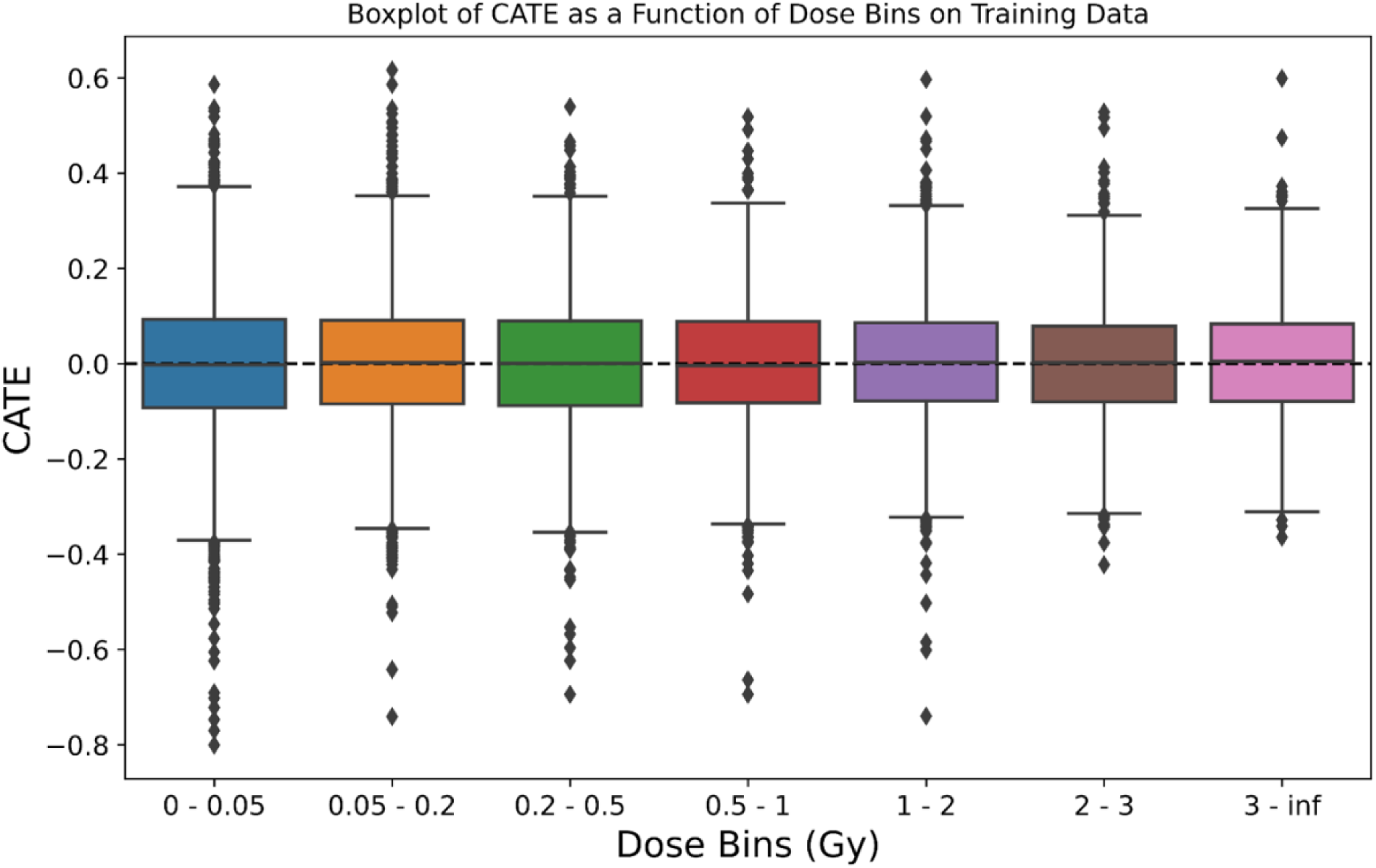
Results of DML analysis of artificially perturbed leukemia data when the outcome variable was replaced with standard normal random numbers. This was intended as a “negative control” for DML reliability.

